# Early real world evidence on the relative SARS-COV-2 vaccine effectiveness of bivalent COVID-19 booster doses: a rapid review

**DOI:** 10.1101/2023.03.28.23287762

**Authors:** M. Sane Schepisi

## Abstract

The objective of this review is to give an overall view of COVID-19 bivalent vaccines knowledge and to explore their early available real world effectiveness evidence in the Omicron era.

Presently, bivalent vaccines are generally offered to all groups eligible for their next booster, as defined by the national vaccination campaign, with varying policies between countries.

The use of bivalent vaccines is supported by immunogenity studies, which, nevetheless, have led to contradictory conclusions, and are not generally designed to measure clinical impact.

In order to critically appraise the available research on real world effectiveness, a systematic literature search was performed: out of 876 references examined, 14 studies were finally included and extracted. The findings of this review demonstrate modest to moderate additional protection of vaccination with bivalent BA.4-5 or BA.1 mRNA-booster vaccines against COVID-19 associated illness and hospitalization, -if compared with having received a monovalent dose as booster-, during a period when BA.5 and other Omicron sublineage viruses predominated globally,

Considering the complexity of the current immunity situation at global level, and the high level of heterogeneity both at study and at review level, these findings must be taken with caution. Further research on SARS-CoV-2 vaccine effectiveness against emerging SARS-CoV-2 variants is encouraged.

## Introduction

The virus that causes COVID-19 changes over time. Monovalent COVID-19 mRNA vaccines were developed against the spike protein of the ancestral SARS-CoV-2 virus and were found to provide cross-reactive immune protection against Alpha and Delta SARS-CoV-2 variants ^1^. The SARS-CoV-2 Omicron variant emerged in November 2021 and diversified into sublineages which were associated with decreased protection from vaccination with monovalent vaccine ^2^.

To address issues of both waning efficacy since the last vaccine dose and the attenuated efficacy of COVID-19 vaccines against variants that escape the immune response directed against spike proteins targeted by the original vaccines, some countries have introduced bivalent mRNA COVID-19 vaccine boosters that encode spike protein from the original SARS-CoV-2 strain and from the Omicron variants. In some countries, the Omicron spike protein encoded in the bivalent mRNA COVID-19 vaccines is based on the BA.1 subvariant; in the United States, it is based on the BA.4 and BA.5 subvariants. Pfizer-BioNTech and Moderna bivalent booster vaccines each contain equal amounts of spike mRNA from the ancestral and Omicron BA.4/BA.5 strains.

The underlying assumption is that using a bivalent versus monovalent vaccine could beccome analogous to updating the seasonal influenza vaccine: new vaccines are produced each year so that the antigenic targets match circulating virus, and these vaccines are made available globally prior to repeated clinical evaluation because of extensive experience with prior versions.

### Immunogenicity studies

Data from immunogenicity studies evaluating bivalent vaccines are mixed and some have not yet been peer reviewed. Postauthorization immunogenicity studies have shown similar neutralizing antibody titers to BA.4/BA.5 after receipt of either a monovalent or BA.4/BA.5–containing bivalent vaccine as a fourth dose ^3 4^. Some studies suggest that bivalent boosters that include the BA.4/5 spike protein induce higher antibody levels against BA.4/BA.5 virus compared with pre-booster levels and compared with monovalent boosters ^5 6 7 8^. Some findings also suggest that the antibody response elicited by the bivalent boosters sufficiently neutralizes other Omicron subvariants, such as BQ.1.1 and XBB. However, in other studies, the antibody response to the bivalent booster was similar to that with the monovalent booster and had minimal neutralizing activity against other Omicron subvariants ^9 10^.

### Current recommendations

The European Medicines Agency (EMA) authorized the first bivalent vaccines in September 2022 ^11 12^. On 6 December 2022 the EMA Emergency Task Force concluded that the bivalent original/Omicron BA.4-5 mRNA vaccines may also be used for primary vaccination ^13^.

On September 1, 2022, the CDC Advisory Committee on Immunization Practices recommended a bivalent COVID-19 mRNA booster dose developed against the spike protein from ancestral SARS-CoV-2 and Omicron BA.4/BA.5 sublineages, for persons who had completed at least a primary COVID-19 vaccination series (with or without monovalent booster doses) ≥2 months earlier ^14^. CDC currently recommeds that all persons aged ≥5 years should receive 1 bivalent mRNA booster dose ≥2 months after completion of any FDA-approved or FDA-authorized monovalent primary series or monovalent booster dose. As a future strategy the FDA has proposed simplifying the Covid-19 vaccination strategy by administering a booster dose in the spring to a smaller group of people, such as the elderly and immunocompromised ^15^, while two annual doses are being considered for immunocompromised adults and children ^16^.

In EU/EEA the bivalent Omicron-adapted vaccines are currently authorised for children aged five years and above for booster vaccination ^17 18^. The majority of EU/EEA countries (Austria, Croatia, Czechia, Denmark, Estonia, Finland, Germany, Hungary, Iceland, Ireland, Lithuania, the Netherlands, Norway, Slovenia, Spain, Sweden) offer the adapted bivalent vaccines to all groups eligible for their next booster ^19^. Recommendations regarding the time intervals between previous infection/vaccination and the second booster varies across European countries, with most countries recommending a waiting period between 3 and 6 months ^20 21 22^. In Italy, the current recommendation is 120 days (4 months)^23^.

Canada’s National Advisory Committee on Immunization published a document entitled “Guidelines on Booster Doses“ on 20 January 2023. entitled “COVID-19 Booster Dose Guidelines: Initial Considerations for 2023“ ^24^. The report mentions different options that will determine the timing of possible booster doses should the health situation warrant it. In Quebec, the “Comité sur l’immunisation du Québec” recommends a booster dose for winter and spring of 2023 to persons considered at high risk who have not yet been infected and whose last booster dose was at least 6 months ago. This recommendation also applies to immunocompromised persons aged 5 years and older, whether or not they have been previously infected ^25^.

In the UK, the Joint Committee on Vaccination and Immunisation (JCVI) recommends an early booster campaign in autumn 2023 for those at high risk of severe severe form. The JCVI is also considering a booster dose in the spring for a smaller group of people, such as the elderly and the immunocompromised ^26^.

In Australia, the Australian Technical Advisory Group on Immunisation (ATAGI) recommends a booster dose before June 2023. a booster dose before June 2023, at least 6 months after the last dose or infection, adults aged 18-64 years ^27^, and children and adolescents aged 5-17 years with co-morbidities and/or disabilities requiring complex care and increasing the risk of severe severe form of Covid-19. ATAGI states that in the coming years, regular administration of vaccine doses, as is done for influenza, will probably be necessary, especially for people at risk of developing severe forms of Covid-19.

The WHO indicated in May 2022 that the available data did not allow for a projection beyond the 2nd booster dose ^28^. In a press conference on 15 February 2023, the European Medicines Agency (EMA)^29^ indicated that it expects vaccination campaigns to take place mainly once a year (administration of a single booster dose each year regardless of previous vaccination status). The EMA assumes that the most appropriate timing for a vaccination campaign against Covid19 is the same as for existing campaigns for other respiratory viruses such as influenza, although it is not clear when the campaign should take place.

To date, little data exist on the effectiveness of the bivalent mRNA-boosters against Covid-19 outcomes. In order to critically appraise the available evidence, published primary real world effectiveness research studies were systematically screened, identified, and synthesised.

## Methods

This review is registered in the PROSPERO International prospective register of systematic reviews (`CRD42023409058`).

A systematic search to identify vaccine effectiveness studies on SARS-COV-2 bivalent vaccines was performed.

### Search criteria

Three different web engines, including early-stage research platforms were used: PubMed, medRxiv, and the Global research on coronavirus disease (COVID-19) database. The following terms were used: “COVID-19”, “vaccine effectiveness”, and “bivalent”. References of eligible studies were screened for inclusion. No restriction on language, setting or publication date was imposed. The search was last updated on the 20th of March 2023.

### Inclusion and exclusion criteria

Eligible articles were real world studies reporting vaccine effectiveness findings on on at least one among the following outcomes: infection based on self-report or presence of anti-nucleoprotein antibodies, hospitalization due to COVID-19 illness as the only outcome, emergency department/urgent care (ED/UC) encounters, death.

As additional outcomes the following were considered: sublineage-specific vaccine effectiveness; vaccine effectiveness waning.

### Selection

Studies were selected through a 2-step procedure: first based on title and abstract and then on full text screening. The RAYYAN web app for systematic reviews was used (https://www.rayyan.ai/). The full process of inclusion is depicted in Figure 1.

**Figure 1.**
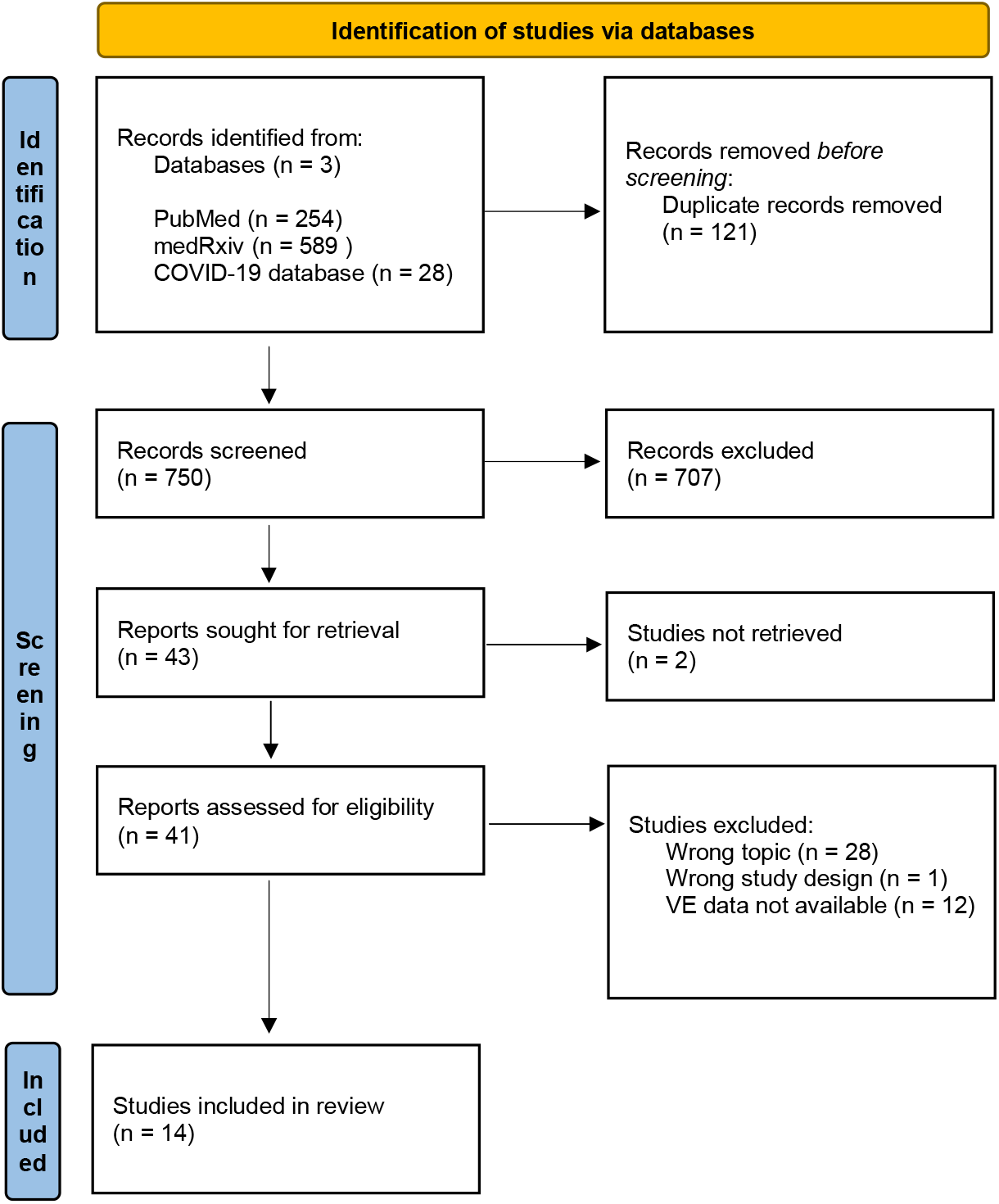
Prisma Flow Diagram - Selection process *From:* Page MJ, McKenzie JE, Bossuyt PM, Boutron I, Hoffmann TC, Mulrow CD, et al. The PRISMA 2020 statement: an updated guideline for reporting systematic reviews. BMJ 2021;372:n71. doi: 10.1136/bmj.n71

### Extraction

A data extraction excel sheet was realised, and the following items were identified and extracted from each included study: Author, Country, Study design, Study period, Study population, Total Number of participants, Number of bivalent vaccine recipients, vaccine type, Age (in years), Number of monovalent booster doses received, Relative Vaccine Effectiveness (VE) in % of self-reported infection, with 95% Confidence Intervals (CI), Relative VE in % of COVID-associated illness (with 95% CI), Relative VE in % of hospitalization (with 95% CI), Relative VE in % of death (with 95% CI). Relative vaccine effectiveness (rVE) data (as point estimates and the corresponding 95% confidence intervals) was extracted from each study, as the benefit of a booster dose of bivalent vaccine compared to the last dose of monovalent vaccine, calculated in percentage points. If not clearly reported in the original studies, rVE was estimated the as 1 minus the corresponding hazard ratio.

### Strategy for data synthesis

If available, sequencing-specific vaccine effectiveness data will be identified, reported and evaluated. Meta-analysis was not planned, due to expected high heterogeneity.

## Results

A total of 876 references were examined and 14 ^30-44^ studies were finally included and extracted.

Most researchers used a retrospective/prospective cohort study design ^30 33 34 35 40 41 43 44^ with a time to event analysis. A matched cohort design was used by a french study ^35^. Some studies had a test-negative case control design, where the odds of having received versus having not received a bivalent booster dose among case-patients (those who received a positive SARS-CoV-2 test result) and among control patients (those who received a negative SARS-CoV-2 test result) were compared ^31 32 36 37 38 39^. The main outcome considered in this review was the relative vaccine effectiveness (rVE) of a bivalent booster dose compared with that of ≥1 monovalent vaccine doses administered. Time since last monovalent dose administered was identified and taken into account in most studies. Most Authors stratified their findings according to time thresholds ^31 32 36 37 40 43.^. One study included only results according to a previusly established minimum period of time elapsed since last monovalent dose ^38^. Few studies reported hospitalization due to COVID-19 illness as the only outcome ^37 38 39^; other studies reported hospitalization data together with data on emergency department/urgent care (ED/UC) encounters ^36^ or on death ^40 41 43 44^; COVID-associated illness was the only outcome for five studies ^31 32 33 34 35^. Link-Gelles et al., in both included studies, considered individuals reporting symptoms consistent with COVID-19 who performed NAAT tests at retail pharmacies ^31 32^. Only one study evaluated self-reported infection ^30^.

Prior SARS-CoV-2 infection based on self-report or presence of anti-nucleoprotein antibodies was evaluated by few studies ^30 33 34 35^, while most Authors collected this information but did not account for it in their analyses ^31 36^.

Main characteristics and findings of included studies are reported in Table 1.

**Table 1.** Main characteristics of 14 included studies. Relative vaccine effectiveness for self-reported infection, COVID-19 associated illness, hospitalization, and death.

|  | Author | Country | Study design | Study period | Study population |  | Relative VE%<br>self-reported<br>infection<br>95% CI | Relative VE%<br>COVID 19-associated<br>illness<br>95% CI | Relative VE%<br>hospitalization<br>95% CI | Relative VE%<br>death<br>95% CI |
| --- | --- | --- | --- | --- | --- | --- | --- | --- | --- | --- |
|  |  |  |  |  | Participants (N)<br>Bivalent booster vaccinated (N)<br>vaccine type | Age (y) |  |  |  |  |
| 1 | AUVIGNE | France | Matched cohort study | 10 Oct. 2022 – 6 Mar. 2023 | 136.852<br>BA.4-5 | ≥60 | - | 8 (0-16) | - | - |
| 2 | ANDERSSON | Denmark, Finland, Norway and Sweden | Cohort study using target trial emulation | 1 Jul. 2022 – 12 Dec. 2022 | 3.368.697<br>1.290.999<br>BA.4-5 (38%), BA.1 (30%) | <50 (Denmark and Sweden)<br><60 (Finland)<br><65 (Norway) | - | - | BA.4-5<br>80,5 (69,5-91,5)<br>BA.1<br>74,0 (68,6-79,4) | BA.4-5<br>77,8 (48,3-100)<br>BA.1<br>80,1 (72,0-88,2) |
| 3 | ARBEL | Israel | Retrospective cohort study | 24 Sept. - 12 Dec. 2022 | 85.314 | ≥65 | - | - | 81 (57-92) | 86 (-4-98) |
| 4 | CHATZILENA | UK | Test-negative case-control | 4 Apr. – 30 Jul. 2022<br>21 Sept. 2022 - 23 Jan. 2023 | 864 admissions (Spring-Summer); 884 admissions (Autumn-Winter)<br>182 cases and 682 controls (Spring-Summer); 152 cases and 732 controls (Autumn-Winter)<br>BA.1 (Comirnaty or Spikevax) | ≥75 | - | - | 46,4 (17,5-54.65) | - |
| 5 | FABIANI | Italy | Retrospective cohort study | 12 Sept. - 12 Dec. 2022 | 11.190.236<br>1.205.353<br>BA.4-5 | ≥60 | - | 58,7 (54.6-62.5) (7-90 d*) | - | - |
| 6 | HUIBERTS | Netherlands | Ongoing prospective cohort study | 26 Sept. - 19 Dec. 2022 | 32.542<br>5.504<br>BA.1 | 18-59<br>60-85 | 18-59 y old<br>31 (18-42)<br>prior Omicron infection 20 (7-40)<br>no prior infection 32 (14-47)<br>prior pre-Omicron infection 44 (13-64)<br>60-85 y old<br>14 (3-24)<br>Prior Omicron infection 6 (30-31) | - | - | - |
| 7 | LIN | USA | Cohort study | 1 Sept - 8 Dec. 2022 | 6.242.259<br>1.070.136 | ≥12 | - | - | all ages<br>33,5 (2,9-62,1) (15-99 d#)<br>≥18 y old<br>32,2 (2,5-60,1) (15-99 d#) | all ages<br>36,9 (12,6-64,3) (15-99 d#)<br>≥18 y old<br>35,4 (11,8-62,1) (15-99 d#) |
|  |  |  |  |  |  |  |  |  | ≥65 y old<br>37,8 (1,0-61,1) (15-99 d#)<br>No prior infection<br>34,7 (6,2-69,9) (15-99 d#) | ≥65 y old<br>41,2 (9,9-71,7) (15-99 d#)<br>No prior infection<br>38,6 (14,8-67,3) (15-99 d#) |
| 8 | LINK-GELLES | USA | Test-negative case-control | 14 Sept. – 11 Nov. 2022 | 360.626 nucleic acid amplification tests performed at<br>9.995 retail pharmacies 5.800 cases and 16.474 controls<br>BA.4-5 | ≥18 | - | 18-49 y old<br>30 (22-37) (2-3 m#)<br>43 (38-48) (4-5 m#)<br>46 (41-50) (6-7 m#)<br>56 (53-58) (≥8 m#)<br>50-64 y old<br>31 (24-38) (2-3 m#)<br>36 (38-41) (4-5 m#)<br>38 (32-43) (6-7 m#)<br>48 (45-51) (≥8 m#)<br>≥65 y old<br>28 (19-35) (2-3 m#)<br>33 (27-39) (4-5 m#)<br>36 (29-41) (6-7 m#)<br>43 (39-46) (≥8 m#) | - | - |
| 9 | LINK-GELLES XBB | USA | Test-negative case-control | Dec 1 2022 –13 Jan. 2023 | 29.175 nucleic acid amplification tests performed at retail pharmacies<br>2.969 cases and 5.289 controls<br>BA.4-BA.5 | ≥18 | - | BA.5<br>18-49 y old<br>52 (48-56) (overall)<br>51 (43-58) (0-1 m*)<br>52 (48-56) (2-3 m*)<br>50-64 y old<br>43 (36-49)<br>54 (43-63) (0-1 m*)<br>39 (30-46) (2-3 m*)<br>≥65 y old<br>37 (28-44) (overall)<br>55 (42-65) (0-1 m*)<br>32 (21-40) (2-3 m*)<br>XBB/XBB.1.5<br>18-49 y old<br>49 (41-55) (overall)<br>50 (36-61) (0-1 m*)<br>48 (39-55) (2-3 m*)<br>50-64 y old<br>40 (28-50) (overall)<br>45 (25-60) (0-1 m*)<br>38 (24-50) (2-3 m*)<br>≥65 y old<br>43 (29-55) (overall)<br>50 (24-68) (0-1 m*)<br>42 (26-54) (2-3 m*) | - | - |
| 10 | POUKKA | Finland | Cohort study | 1 Sept. 2022 - 31 Jan. 2023 | 1.197.700 elderly<br>627.378<br>BA.1 or BA.4-BA.5 | 65-120<br>18-64 | - | - | Elderly<br>67 (43-67) (14-30 d*)<br>67 (43-67) (31-60 d*) | Elderly<br>61 (43-74) (14-30 d*)<br>43 (23-58) (31-60 d*) |
|  |  |  |  |  | 444.683 chronically ill<br>66.871<br>BA.1 or BA.4-BA.5 |  |  |  | 26 (-9-50) (61-90 d*)<br>Chronically ill<br>18 (-1,07-68) (14-30 d*)<br>-43 (-1,07-22) (31-60 d*) | 26 (-13-51) (61-90 d*) |
| 11 | **PUBLIC HEALTH ENGLAND** | UK | Test-negative case-control | 5 Sept. – 25 Dec., 2022 | 9.981 hospitalized patients<br>8617<br>BA.1 or BA.4-BA.5 | ≥50 | - | - | **Corminaty**<br>43,1 (32,3–52,3) (2-4 w*)<br>50,8 (41,5–58,6) (5-9 w*)<br>46,4 (20,1–64,1) (10+ w*)<br>**Spikevax**<br>57,8 (51,2–63,5) (2-4 w*)<br>50,5 (44,1–56,1) (5-9 w*)<br>47,5 (38,5–55,2) (10+ w*)<br>BQ.1<br>**Any bivalent**<br>52,1 (40,2–61,6)<br>**Corminaty**<br>54,1 (36,0–67,1)<br>**Spikevax**<br>51,0 (37,7–61,4)<br>BA.5<br>**Any bivalent**<br>63,6 (53,8–71,3)<br>**Corminaty**<br>53,7 (28,5–70)<br>**Spikevax**<br>65,8 (55,5–73,7) | - |
| 12 | **SHRESTA** | USA | Retrospective cohort study | 12 Sept. 2022 – 21 Feb., 2023 | 48.141<br>12.789<br>Corminaty (87%) or Spikevax BA.4–BA.5 | ≥18 | - | 29 (20-37)<br>BA.4/5 dominant phase<br>19 (5-30)<br>BQ dominant phase<br>4.5 (18-22)<br>XBB dominant phase | - | - |
| 13 | **SURIE** | USA | Test-negative case-control | Sept. 8 – Nov. 30, 2022 | 798 individuals, 74% of whom with multiple underlying conditions<br>20 cases and 59 controls<br>Corminaty or Spikevax BA.4–BA.5 | ≥65 | - | - | 73 (52–85) (2-5m#)<br>78 (57-89) (6-11m#)<br>83 (63–92) (≥12 m#) |  |
| 14 | **TENFORDE** | USA | Test-negative case-control | Sept. 13 – Nov. 18, 2022 | 78.303 ED/UC encounters with COVID-19-like illness<br>3.658 cases and 247 controls<br>Corminaty or Spikevax<br>BA.4–BA.5<br>15.527 hospitalizations with COVID-19-like illness<br>734 cases and 49 controls Corminaty or Spikevax BA.4–BA.5 | ≥18 | - | 31 (19–41) (2–4 m#)<br>42 (32–50) (5–7 m#)<br>53 (43–57) (8–10 m#)<br>50(43–57) (≥11 m#) | 38 (13–56) (5–7 m#)<br>42 (19–58) (8–10 m#)<br>45 (25–60) (≥11 m#) | - |

### Self-reported infection

A study conducted in the Netherlands^30^ used data of 32.542 prospective cohort study participants who previously received primary and one or two monovalent booster COVID-19 vaccinations. Between 26 September and 19 December 2022, the relative effectiveness of bivalent original/Omicron BA.1 vaccination against self-reported Omicron SARS-CoV-2 infection was 31% in 18–59-year-olds and 14% in 60–85-year-olds, adjusted for infection hystory. Prior Omicron infection provided higher protection than bivalent vaccination among persons without prior infection, even though the time since prior Omicron infection was longer than the time since bivalent vaccination.

### COVID-associated illness

The first published estimates of VE for newly authorized bivalent mRNA booster vaccines among over 250,000 symptomatic adults who received testing for SARS-CoV-2 infection at pharmacies nationwide during September 14–November 11, 2022 in USA ^31^ provided initial evidence on the efficacy of bivalent mRNA vaccines in preventing symptomatic SARS-CoV-2 infections compared with previous vaccination with 2, 3, or 4 monovalent vaccines alone. The relative efficacy of a booster dose with the bivalent vaccine was 28 to 31 percent among those who had last been vaccinated two to three months previously and 43 to 56 percent among those last vaccinated more than eight months previously. The results suggest, therefore, that the relative gain in efficacy of a booster dose with the bivalent vaccine compared to the last dose of monovalent vaccine is greater the longer the the time since the last dose of vaccine increases ^31^.

VE estimates for XBB/XBB.1.5 sublineage and for BA.5 sublineage–related symptomatic SARS-CoV-2 infection were obtained by another study from from USA national pharmacy testing during December 1, 2022–January 13, 2023. This report provides the first estimates of bivalent mRNA COVID-19 VE against with XBB-related sublineages. These preliminary estimates showed relative bivalent booster dose VE (compared with 2–4 monovalent doses) to be similar for XBB/XBB.1.5 sublineage– related infections and BA.5 sublineage–related infections. Across age groups, VE was generally similar against BA.5-related infections and XBB/XBB.1.5-related infections. VE against symptomatic BA.5-related infection was 52% among persons aged 18–49 years, 43% among persons aged 50–64, and 37% among those aged ≥65 years ^32^.

A retrospective study (in preprint) conducted by Shrestha et al. at ^33^ the Cleveland Clinic Health System in the USA, including 51.011employees of the centre, aimed to evaluate the efficacy of the bivalent vaccine (original/Omicron BA.4-5) and found an estimated vaccine effectiveness (VE) of 29% (95% C.I., 20%-37%), 19% (95% C.I., 5%-30%), and 4.5% (95% C.I., -18%-22%), during the BA.4/5, BQ, and XBB dominant phases, respectively. Risk of COVID-19 also increased with time since most recent prior COVID-19 episode and with the number of vaccine doses previously received ^33^.

In a recently published Italian retrospective cohort study ^34^ the effectiveness against severe COVID-19 of a second booster dose of the bivalent (original/BA.4–5) mRNA vaccine 7–90 days post-administration, relative to a first booster dose of an mRNA vaccine received ≥ 120 days earlier, was 60% both in persons ≥ 60 years never infected and in those infected > 6 months before. Relative effectiveness in those infected 4–6 months earlier indicated no significant additional protection (10%; 95% CI: −44 to 44) ^34^.

A French matched cohort study recently available as preprint ^35^ included 136.852 individuals of ≥60 years old who received a booster dose between 03/10/2022 and 06/11/2022, when both the bivalent and monovalent vaccines were used in France. Those who received a booster dose with (1) a monovalent Original mRNA vaccine (Pfizer-BioNTech or Moderna) or (2) the bivalent Pfizer-BioNTech Original/BA.4-5 vaccine were matched. After a median follow-up period of 77 days the bivalent vaccine conferred an additional protection of 8% [95% CI: 0% - 16%, p=0.045] against symptomatic SARS-CoV-2 infection compared to the monovalent vaccines. Previous infection was classified according to the dominant variants in France at the time of infection: Delta/PreDelta, Transition from Delta to BA.1, Omicron BA.1 (including the transition between BA.1 and BA.2) and Omicron BA.2 BA.4/5. A recent previous infection with BA.2 or BA.4-5 conferred an additional protection of 74% [95% CI: 78% - 69%] while older previous infections, caused by variants more distinct from the circulating strains, conferred lower but still detectable protection in boosted participants.

### COVID-associated hospitalization

A US study conducted by Tenforde et al. ^36^ in immunocompetent adults aged 18 years and older found that the relative benefit of a booster dose of bivalent vaccine compared to the last dose of monovalent vaccine increased as the time since last vaccination increased. Bivalent vaccines administered after ≥2 monovalent doses were effective in preventing admission for Covid-19 with a relative VE of 31% when the last dose of monovalent vaccine was administered 2-4 months earlier, and of 50% if the last dose of vaccine was administered 11 months or more earlier. The relative EV for hospitalisations was 38% given compared to a last dose given 5-7 months earlier and 45% when the last dose was given 11 months or more ago.

Another US CDC report ^37^ evaluated the bivalent booster’s VE in preventing Covid-19-related hospital admissions in people aged 65 years and older and found high protection of the bivalent BA.4-5 boosters received after ≥2 monovalent mRNA vaccines against Covid19 hospitalization when compared with past (≥2 months) monovalent mRNA vaccination only. The relative VE of the bivalent booster increased with time since last vaccination, regardless of the number of booster doses received: it was 73% when the last booster was 2 months or more old, 78% when the last booster was 6 to 11 months old and 83% when the last booster was 12 months old or more.

In an English analysis ^38^ bivalent boosters with either Pfizer BioNTech (Original/Omicron BA.1) or a Moderna bivalent (bivalent Original/Omicron vaccine) targeting both the ancestral strain and Omicron BA.1 were offered to those in clinical risk groups and those aged 50 years and older from September 2022. VE of the bivalent boosters was estimated against hospitalisation in the period following 5 September 2022 against all Omicron sub-lineages in circulation at the time. Only individuals who had received at least 2 COVID-19 vaccines before 5 September 2022 and with the last of these doses at least 6 months prior to sample date were included in analysis. The incremental protection conferred by the bivalent vaccines estimated relative to those with waned immunity was 43,1% for Pfizer after 2 weeks, and 57,8% for Moderna. Effectiveness remained high at 10 or more weeks after vaccination at 46.4% for the Pfizer booster and 47,5% for the Moderna booster. Moreover VE against hospitalisation for BQ.1 and BA.5 was estimated during a period of co-circulation:. cases were classified as BA.5 or BQ.1 based on sequencing information. The effectiveness of the bivalent booster against hospitalisation with BQ.1 was 52.1% as compared to 63.6% with BA.5, at 2 or more weeks after receiving the booster.

Another very recent UK study, preliminarly available as a preprint ^39^ assessed rVE against hospitalisation for the Spring-Summer (fourth vs third monovalent mRNA vaccine doses) and Autumn-Winter (fifth BA.1/ancestral bivalent vs fourth monovalent mRNA vaccine dose) boosters. This prospective single-centre test-negative case-control study was conducted among ≥75 year-olds hospitalised with COVID-19 or other acute respiratory disease and concluded that bivalent mRNA boosters offer equivalent protection against hospitalisation with Omicron infection to monovalent mRNA boosters given earlier in the year: a monovalent mRNA COVID-19 vaccine as fourth dose showed rVE 46·9% (95% confidence interval [CI] 14·4-67·3) versus those not boosted while a bivalent mRNA COVID-19 vaccine as fifth dose had rVE 46·4% (95%CI 17·5-65), compared to a fourth monovalent mRNA COVID-19 vaccine dose.

### COVID-associated hospitalization and death

Lin et al. ^40^., in a study conducted in the USA, evaluated the effectiveness of a booster dose with a bivalent vaccine compared to a booster dose with a monovalent vaccine. This study was based on data collected over 99 days during which bivalent boosters were administered from 1 September to 8 December 2022, and the previous 99 days during which monovalent booster shots were administered, i.e. from May 25 to August 31, 2022. The results showed that the efficacy of the booster with bivalent vaccine against hospital admissions between days 15 and 99 after administration was 58.7% (95% CI: 43,7%-69,8%) compared to 25,2% (95% CI: -0,2% to 44,2%) for a monovalent vaccine booster. In addition, the efficacy of the bivalent vaccine booster with respect to hospitalisations or deaths was 61.8% (95% CI: 48.2%-71,8%) versus 24,9% (95% CI: 1,4-42,8) with the monovalent vaccine booster.

A wider comparative effectiveness cohort study conducted in Denmark, Finland, Norway and Sweden ^41^ using target trial emulation compared receipt of a bivalent BA.4-5 booster as a fourth dose with having received three vaccine doses with the AZD1222, BNT162b2 and/or mRNA-1273 vaccines. The efficacy of the second booster with the bivalent BA.4-5 vaccine with respect to hospitalizations compared to no booster after the first booster (3rd dose with a monovalent vaccine) was 80,5% (95% CI: 69,5%-91,5%) and 74% (95% CI: 68,6%-79,4%) with the bivalent BA1 vaccine. With regard to deaths, the effectiveness of the second booster was 77,8% (95% CI: 48,3%-100%) for the bivalent BA.4-5 vaccine and 80,1% (95% CI: 72%-88,2%) for the bivalent BA.1 vaccine compared to the first booster.

Follow-up data reported by the CDC in the United States ^42^ on the surveillance of COVID-19-related infections and deaths by vaccination status (13 September to 23 October 2022 for bivalent vaccine), showed that the risk of death was lower in those who received a booster dose with a bivalent vaccine compared to those boostered with monovalent vaccine. with a monovalent vaccine.

A recent study preliminarily published in medRxiv and conducted in Finland ^43^[POUKKA] involved 1.197.700 individuals aged between 65 and 120 years and 444.683 chronically sick people aged between 18 and 64 years. In Finnish register-based cohort analyses, the risk of severe COVID-19 outcomes among those who received bivalent vaccination between 1 September 2022 and 31 January 2023 was compared to those who did not. Among elderly aged 65–120 years, bivalent vaccination reduced the risk of hospitalisation and death due to COVID-19. Among the elderly the hazard ratios comparing exposed and unexposed ranged from 0,36 to 0,43 during the first 14–30 days since bivalent vaccination but signs of waning were observed as soon as two months after vaccination. Among the chronically ill aged 18–64 years bivalent vaccination did not reduce the risk of severe COVID-19 outcomes.

A similar Israeli ^44^ retrospective cohort study included more than 600.000 people aged over 65 years among which over 85.000 participants received the booster with the bivalent vaccine. In this study slightly higher BA.4-5 bivalent booster effectiveness against severe COVID-19 outcomes was found: VE was 81% against hospitalisations and 86% against Covid-19-related deaths (although the estimate for Covid-19 death was statistically insignificant).

## Discussion

The findings of this review show modest to moderate protection of vaccination with bivalent BA.4-5 or BA.1 mRNA-booster vaccines as a fourth dose against COVID-19 associated illness and hospitalization during a period when BA.5 and other Omicron sublineage viruses predominated globally, if compared with having received two or more monovalent vaccine doses. Bivalent vaccines restore protection observed to wane after monovalent vaccine receipt, as demonstrated by increased relative VE with longer time since the most recent monovalent dose. Nevertheless the added benefit in preventing SARS-CoV-2 Omicron infection seems small, especially in persons with prior Omicron infection.

The majority of included studies did not assesss the presence of the circulating strains represented in the bivalent vaccines. A significant protective effect was not found when the XBB lineages were dominant, and protection if any was likely to be clinically insignificant ^33^. VE against hospitalisation for BQ.1 and BA.5 was estimated during a period of co-circulation by Public Health England researchers, based on sequencing information, and effectiveness point estimate were lower for BQ.1 ^38^. Only one study ^32^ reported updated data on the XBB/XBB.1.5 sublineages which are gaining predominance worldwide.

Among the chronically ill a Finnish study ^43^ did not observe bivalent vaccination to reduce the risk of severe COVID-19 outcomes, although another study found a benefit on working-age adults ^37^.

According to one wide cohort study ^41^ bivalent BA.4-5 boosters conferred moderately greater vaccine effectiveness against Covid-19 hospitalization compared with bivalent BA.1 boosters.

Hybrid immunity was investigated by few studies ^34 35^ which confirmed that additional protection was acquired by recent infections while lower but still detectable protection in boosted participants originated from older infections. According to the findings reported by Huiberts et al. prior Omicron infection provided higher protection than bivalent vaccination among persons without prior infection, even though the time since prior Omicron infection was longer than the time since bivalent vaccination ^30^.

The findings of this review are in line with those of an ecologic study by Johnson et. al ^45^ conducted from October 2021 to December 2022, where overall incidence rates among unvaccinated persons were compared to rates in persons with only monovalent doses or bivalent boosters. Receipt of bivalent booster added protection against infection for circulating Omicron BA.4/BA.5 sublineages, and evidence of waning protection against infection 3 months after bivalent booster dose receipt was observed. Regarding death, comparisons during the late BA.4/BA.5 period of monovalent and bivalent boosters found that bivalent boosters restored protection against mortality.

A number of limitations at study level have been identified.

Generally, low numbers in bivalent vaccine uptake may have caused unprecise estimates.

The comparison between bivalent and monovalent vaccines, due to changing authorization rules, was not performed during the same period, and the persons vaccinated earlier were necessarely and systematically different from those vaccinated later. Moreover, these early studies could not assess the long-term durability of bivalent booster vaccination protection because of the short period of observation since bivalent dose receipt.

Regarding outcome detection, a substantial proportion of the population may have had prior unrecognized asymptomatic Omicron variant infection, masking the protective effect of the bivalent dose due to natural immunity, and generating a weaker than expected vaccine effectiveness. Most studies did not account for previous SARS-CoV-2 infections in their analyses. The underestimation of bivalent vaccine effectiveness may have also been caused by a good baseline protection due to monovalent vaccination.

Testing behaviour may have differed between the exposed and the unexposed groups, as those who received the bivalent vaccine would have been less inclined to get tested for the same symptoms after getting the bivalent vaccine than before, providing greater opportunity to detect infection in the non-vaccinated than the bivalent vaccinated state, thereby having the effect of overestimating vaccine effectiveness.

Due to the current clinical characteristics of COVID-19 associated illness, there were too few severe illnesses for the study to be able to determine if the vaccine decreased severity of illness.

With regards to COVID-19 deaths, they reported as related because they ocurred in hospitalized accidentaly found to be SARS-CoV-2 positive cases.

Finally, most included studies were conducted prior to the emergence of the current predominating omicron subvariants BA.4 and BA.5 and sublineage-specific VE were not estimated.

The main limitation at review level was the heterogeneity of vaccines administered and, more importantly, in time elapsed from the last monovalent vaccine dose.

## Conclusion

Early data from real world studies on bivalent vaccines seem to support the effectiveness findings reported for monovalent vaccines. The interpretation of these results is skewed by the complicated current heteorogeneous situation of our immunity, the lower than predicted coverage of bivalent vaccination, and the shifting dynamics of Omicron sublineages balance and composition.

With the circulation of sublineages of the BA.4/BA.5 Omicron variants like BQ.1 and BQ.1.1, ongoing monitoring is required to assess the longevity of the added protection. In order to determine the best timing for receiving bivalent vaccine booster doses and to develop vaccination plans for the foreseeable future, it is especially important to evaluate VE against outcomes like COVID-19-associated severe respiratory illness or death.

It is especially crucial to prevent diseases that require medical attention and to lessen the burden on the healthcare system when several respiratory viruses are co-circulating. Long-term planning for Covid-19 immunization campaigns should include a variety of considerations, including epidemiological patterns, potential seasonality, impact on health systems, and economic concerns, in addition to the introduction of novel variants and the efficacy of new variant-adapted vaccines.

## Data Availability

All data produced in the present study are available upon reasonable request to the author.

